# Rethinking HIV care for youth: Insights from qualitative research with youth in Chad

**DOI:** 10.1101/2024.08.20.24312037

**Authors:** Esias Bedingar, Ferdinan Paningar, Ngarossorang Bedingar, Eric Mbaidoum, Naortangar Ngaradoum, Rifat Atun, Aisha K. Yousafzai

**Affiliations:** Global Health and Population Department, Harvard T.H. Chan School of Public Health, Boston, MA 02215, USA; Alma, Centre de Recherche en Systèmes de Santé, Porte 107 Chagoua, N’Djamena, Chad; Bucofore, Quartier Béguinage, Rue Joseph Brahim Seid, N’Djamena, Chad; Croix Bleue Tchadienne, Porte N128, 7eme arrondissement, Chagoua, N’Djamena, Chad; Réseau National des Personnes Vivants avec le VIH, N’Djamena, Chad

## Abstract

Youth ages 15-24 years are significantly impacted by the HIV/AIDS epidemic, representing approximately 37% of new infections globally. This demographic is especially vulnerable in sub-Saharan Africa, where over 80% of HIV-positive youth reside. In Chad, youth face significant barriers to effective HIV care, including high prevalence rates, particularly among young women, and substantial disparities across regions. Despite overall reductions in new HIV infections, youth remain disproportionately affected, necessitating targeted interventions to improve HIV care outcomes. This study employed thematic analysis to conduct a secondary data analysis of previously collected qualitative data. The analysis focused on understanding the pathways to care for youth living with HIV (YLHIV) in Chad, from diagnosis to antiretroviral therapy (ART) adherence. Data were collected through focus group discussions with 52 youth and 48 service providers, including healthcare workers and community actors. Grounded theory analysis was used to identify barriers and facilitators to HIV care, with data transcribed, translated, and analyzed assisted with ATLAS.ti software (Version 7.6.3). Youth reported significant barriers, including financial constraints, logistical challenges, and fear of stigma. Facilitators included supportive healthcare providers, peer support, and specialized HIV care facilities. Psychological barriers and lack of awareness were also critical factors influencing HIV testing and care engagement. The study highlighted the importance of tailored, youth-friendly services to improve care outcomes. To address the unique challenges faced by youth in Chad, healthcare services must be redesigned with active youth participation to be more adolescent-friendly and supportive. Interventions should focus on reducing stigma, improving accessibility, and providing continuous psychosocial support. By addressing these barriers, it is possible to enhance the HIV care continuum for youth, ultimately improving their health outcomes and contributing to the broader goal of ending the HIV epidemic.

## Introduction

Youth (ages 15-24) are a crucial demographic in the global fight against HIV/AIDS, comprising about 37% of new HIV infections worldwide [1–3]. Adolescents and young people (10-24 years) are especially vulnerable, experiencing high HIV prevalence and suboptimal treatment outcomes [4, 5]. This issue is pressing in sub-Saharan Africa, where over 80% of youth living with HIV (YLHIV) reside [6–8], with high rates of HIV-related mortality among young people [9]. In Chad, the HIV prevalence was 1.0% in 2022, down from 1.6% in 2010 and 1.4% in 2015, but varies significantly across provinces [8, 10].

In Chad, young women face higher HIV infection rates compared to young men due to the “triple threat” of new HIV infections, sexual and gender-based violence, and adolescent pregnancies [11]. In 2022, 26.3% of new HIV infections in Chad were among youth aged 15-24 [8]. Despite a 48% overall decrease in new HIV infections since 2010, youth remain disproportionately affected [8], highlighting a gap in prevention and care services tailored to this group [12].

Youth face numerous barriers in the HIV care continuum, from testing to achieving viral suppression, including stigma, transportation issues, and the transition from pediatric to adult care [13, 14]. These challenges lead to lower linkage to care, higher attrition, and poor adherence to ART among youth compared to other age groups [15, 16]. Limited studies indicate poorer outcomes for HIV-infected youth in the care pathway [17], with significantly lower retention rates in care before initiating ART and low adherence even after starting treatment [18].

Service delivery interventions such as counseling, peer support, financial incentives, and adolescent-friendly clinics show promise in improving linkage, retention, and adherence among youth [17, 19, 20]. Integrating biomedical prevention with behavioral, psychosocial, and structural approaches can enhance the HIV care continuum, especially in the context of Chad’s complex sociocultural landscape.

This study aims to examine the pathways to care for youth to improve linkages from HIV diagnosis to ART initiation, retention, and adherence. By understanding the challenges youth face, we aim to identify strategies to enhance the continuum of HIV care for this vulnerable population.

## Methods

### Study design

This study is a secondary data analysis from a parent study employing grounded theory design [21, 22]. The stages and processes employed for the parent study are shown in the Supporting Information file (S1 File). The original data were collected to understand youth’s sensemaking and service utilization in the context of SRH and HIV care in Chad.

### Sampling

Participants were selected using purposeful criterion sampling to ensure a diverse and relevant sample of youth and healthcare providers. The sampling frame was bound to the province of N’Djamena to maintain contextual relevance while allowing for a variety of perspectives. Youth, aged 15-24 years, were recruited and divided into two groups: those living with HIV and those without HIV. The final sample included 52 youth, with an almost equal distribution of gender and HIV status. For adolescents living with HIV, we collaborated with the National Executive Secretary of the Chadian National Network of Associations of People Living with HIV (RNTAP^+^) who helped identify those willing and eligible for the study. Eligibility was defined as either a male or female ages 15-24 years living with HIV. From the final list provided, we randomly selected adolescent participants.

The study also included 48 service providers, divided into 3 focus group discussions (FGDs), including nurses, doctors, and community actors to provide comprehensive insights into the healthcare environment. Each group of service providers was sub-divided into 3 groups of 4 people for healthcare workers, and 4 groups of 6 people for community actors. For both nurses and doctors, the groups reflected health facility type, including public and specialized facilities for SRH and HIV care. Eligibility criteria for health facilities included the existence of pediatric, maternity and SRH service areas, as well as being frequently visited by youth. After selecting these facilities, the second step consisted of obtaining a list of nurses and doctors who had been working at the facility for at least 6 months to ensure sufficient exposure to youth. For community actors, the four groups included those existing in Chad such as peer educators, mentor moms, expert patients, and psycho-social counselors [23].

### Data collection

Data were collected through FGDs conducted from 24/03/2024 to 27/03/2024. The FGDs were designed to elicit detailed accounts of participants’ experiences, beliefs, and behaviors related to HIV care and ART. The sample of participants needed to encompass both homogeneity and heterogeneity at various levels. For instance, homogeneity was achieved through bounding the sampling to one province, as well as keeping sample and facility characteristics close to each other (e.g., narrowing age, YLHIV, youth living with no HIV). On the other hand, heterogeneity was achieved through increasing the number of FGD participants and bracketing participants by age groups. Typical FGDs include 6-10 participants, but since our study had participatory activities, we designed it to have 16 youth in each group, and further sub-divided them into 4 groups of 4 participants for the activities (S2 File). After data collection, this resulted in a total sample size of 52 youth, stratified by gender. Service providers participated in FGDs, discussing their roles, perceptions of adolescent-friendly healthcare, and the challenges they face in delivering HIV care to youth. Participatory action research (PAR) is a good methodological framework for social inclusion [24]. For that reason, youth and service providers took part in open-ended questions for each theme, as well as participatory activities for decision-making, community mapping, and sorting influential factors for stages 2 and 3 (S1 File). For data collection, separate topic guides were used for adolescents, healthcare workers, and community actors (S3-S5 Files).

The data collection process was facilitated by fourteen locally trained data collectors, divided into interviewers and note-takers to conduct the FGDs and participatory activities. However, study visits with female study participants occurred with at least one female study member in the room, and male participant study visits occurred with at least one male study team member in the room. FGDs were conducted in French and Arabic in a private setting. All FGDs sessions, lasting between 90-120 minutes, were audio-recorded with participant consent, transcribed verbatim, and imported into ATLAS.ti (version 7.6.3) to manage data and assist the analysis process [25]. To ensure transcript quality, a rigorous quality assurance process was implemented: audio recordings were cross-checked against transcripts by a secondary team member for accuracy, and the first author reviewed a sample of transcripts for consistency. Data collectors underwent comprehensive training and calibration to standardize data collection techniques, and iterative feedback loops were established to address and correct any transcription issues identified during regular debriefing sessions.

### Data analysis

We conducted a thematic analysis using both deductive and inductive approaches [26]. Thematic analysis was chosen for its flexibility and ability to identify, analyze, and report patterns (themes) within the data, providing a rich and detailed account of the participants’ experiences and perceptions. For deductive coding, the longitudinal HIV care continuum, a public health model, was used to guide coding (deductive codes) and organize themes [27]. The choice of these deductive themes, including HIV testing, linkage to HIV care and ART, and retention in ART care, were justified by their fundamental role in the HIV care continuum and their relevance to the experiences and challenges described by youth in the transcripts [17, 27]. These themes provided a structured approach to analyzing the key issues in HIV prevention and treatment among young people.

The process of thematic analysis followed the Braun and Clarke’s six-phase method to ensure a thorough and rigorous examination of the data [28]. Initially, we familiarized ourselves with the data by reading and re-reading the transcripts, making initial notes and observations. This was followed by generating initial codes from the data in a systematic fashion across the entire dataset, which involved identifying significant features related to the research questions. These codes were then collated into potential themes, grouping related codes to reflect broader patterns of meaning. Next, we reviewed and refined these themes to ensure they accurately represented the data, checking if they worked in relation to the coded extracts and the entire dataset. Once themes were clearly defined, we named and described each theme, outlining their scope and focus. Finally, we produced a report that included detailed analysis of selected extracts, illustrating the themes and their relevance to the research questions. See Supporting File (S1 Table) for an overview of the themes and subthemes with sample supporting quotes.

To maintain reliability and integrity of the analysis, we conducted the thematic analysis in the original languages of the transcripts, French and Arabic, before translating the results into English. This approach ensured that linguistic and cultural nuances were preserved and accurately reflected in the final analysis.

Two care maps were created to represent the current pathways to care by HIV status (positive and negative) from the participatory group activity in the youth FGD, which explored decision making processes for participation (or not) in HIV services [29]. Figures 2A and 2B represent an aggregate of each FGD’s care maps for HIV negative and positive youth, respectively. Within each figure, tables describe distinct healthcare resources at each step of care (S2-S3 Tables).

### Research trustworthiness

To ensure the trustworthiness of the research, we adhered to the evaluative standards set forth by Lincoln and Guba (1985), which include credibility, transferability, dependability, and confirmability [30]. Credibility was established through prolonged engagement with the data and participant verification. The involvement of locally trained data collectors who were familiar with the cultural context further enhanced the credibility of the data collection process. Transferability was facilitated by providing thick descriptions of the study context, participant demographics, and data collection methods, allowing readers to assess the applicability of the findings to other contexts. Dependability was achieved by maintaining a detailed audit trail of the research process, including the steps taken during data collection, coding, and thematic analysis. Finally, confirmability was ensured through triangulation, peer debriefing, and maintaining a reflexive journal to document potential biases and how they were managed throughout the research process. Additionally, saturation was achieved at *n = 12* for youth and *n = 9* for service providers, as no new themes or insights emerged from the data. These measures collectively ensured that the findings are trustworthy and accurately represent the experiences of the participants.

### Reflexivity

Reflexivity was a critical component of this study, given the complex sociocultural context of Chad. The research team, comprising individuals from diverse cultural and disciplinary backgrounds, engaged in continuous self-reflection to recognize and mitigate potential biases. As the Principal Investigator and a native of Chad, EB (first author), brought essential local knowledge and cultural sensitivity to the study, particularly regarding the complexity of Chad’s healthcare system and socio-economic landscape. Additionally, the collaboration with the RNTAP^+^, composed entirely of Chadian nationals familiar with the local context and HIV, further ensured that the study was deeply rooted in the local reality. The team also included international researchers with extensive expertise in qualitative and HIV research, which guaranteed methodological rigor. The inclusion of locally trained interviewers who were proficient in the local languages and familiar with cultural nuances was essential in capturing authentic data. Regular team meetings were held to discuss and reflect on the data collection process, emerging themes, and any potential biases. The use of a reflexive journal allowed researchers to document their thoughts, decisions, and interactions with participants, which helped in maintaining an awareness of how their perspectives might influence the research. This reflexive practice ensured that the findings were grounded in the participants’ realities rather than the researchers’ preconceptions.

### Ethics approval

Ethical considerations were paramount in this study, given the sensitive nature of the topic and the vulnerability of the adolescent participants. Ethical approval was obtained from both the Harvard T.H. Chan School of Public Health’s Institutional Review Board (IRB) protocol #IRB23-1743 and the National Committee on Bioethics of Chad (#010/MESRS/SE/SG/2024). Informed consent was obtained from all participants, with additional assent from guardians for participants who were younger than 18 years of age. The consent process involved reading the consent forms aloud and allowing participants to ask questions, ensuring they fully understood their participation rights and the study’s purpose. To maintain confidentiality, pseudonyms were used, and all identifying information was removed from the transcripts. Data were securely stored, and only the research team had access to them. Additionally, the research design incorporated measures to minimize potential distress, such as the presence of same-gender interviewers for sensitive topics and providing participants with contact information for counseling services if needed. These ethical practices ensured the protection and well-being of the participants throughout the study.

## Results

### Study characteristics

Table 1 presents the demographics of the participants. A total of 23 focus group discussions were conducted with youth and service providers, including healthcare workers and community actors. Among youth, the study involved 23 HIV-positive and 29 HIV-negative participants. In addition, 48 service providers were divided into 3 groups, including 12 doctors, 12 nurses, and 24 community actors, mostly coming from public health facilities (79.2%). The findings consisted of 3 main themes and 10 subthemes: (1) HIV testing services, (2) linkage to HIV care and ART, and (3) retention on ART (Table 2).

**Table 1.**
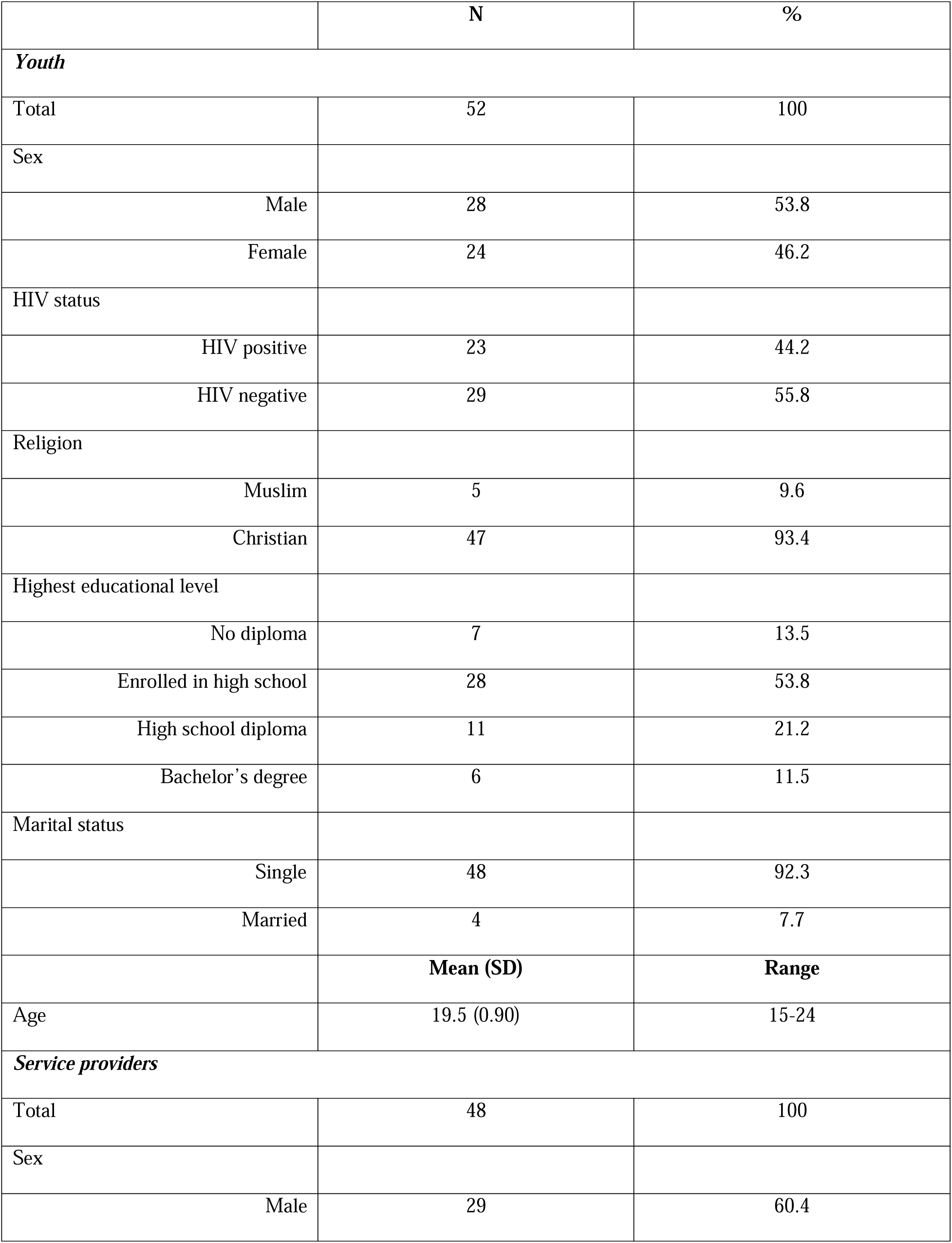
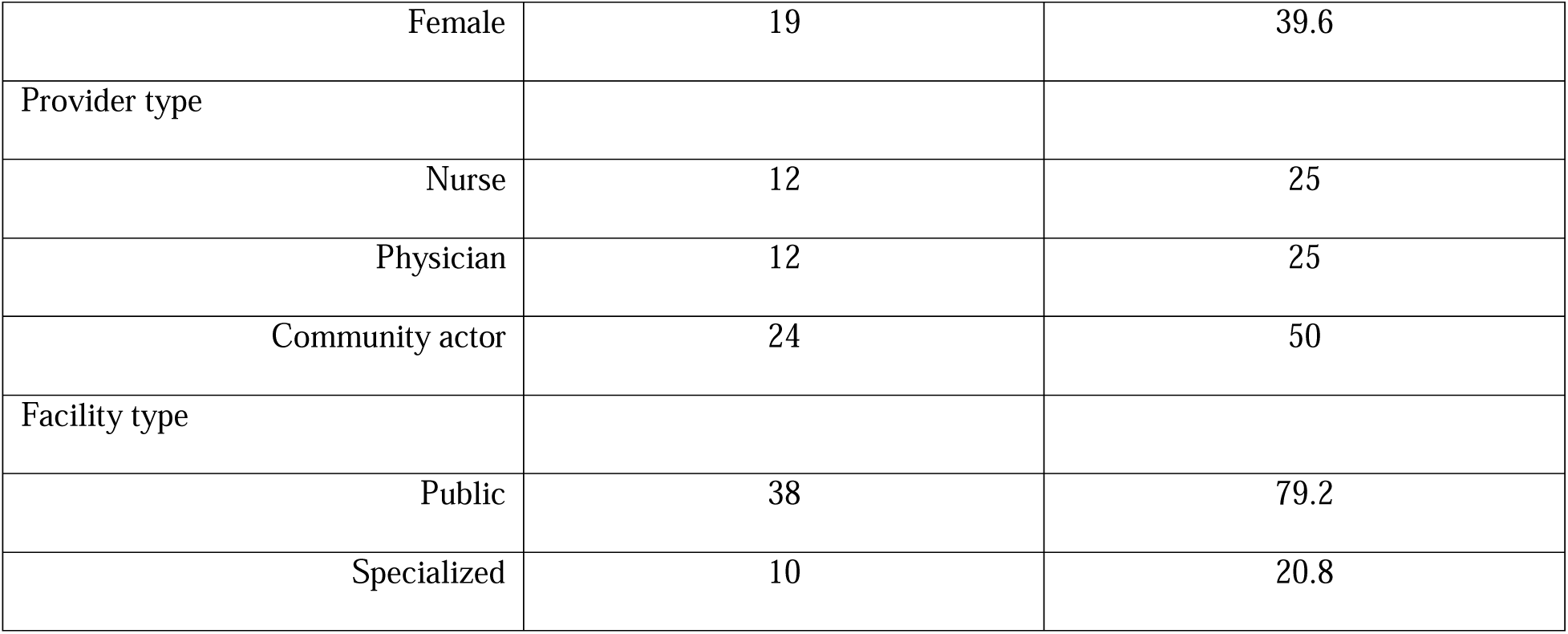
Sample demographics for youth and service providers.

|  | N | % |
| --- | --- | --- |
| ***Youth*** |  |  |
| Total | 52 | 100 |
| Sex |  |  |
| Male | 28 | 53.8 |
| Female | 24 | 46.2 |
| HIV status |  |  |
| HIV positive | 23 | 44.2 |
| HIV negative | 29 | 55.8 |
| Religion |  |  |
| Muslim | 5 | 9.6 |
| Christian | 47 | 93.4 |
| Highest educational level |  |  |
| No diploma | 7 | 13.5 |
| Enrolled in high school | 28 | 53.8 |
| High school diploma | 11 | 21.2 |
| Bachelor's degree | 6 | 11.5 |
| Marital status |  |  |
| Single | 48 | 92.3 |
| Married | 4 | 7.7 |
|  | **Mean (SD)** | **Range** |
| Age | 19.5 (0.90) | 15-24 |
| ***Service providers*** |  |  |
| Total | 48 | 100 |
| Sex |  |  |
| Male | 29 | 60.4 |
| Female | 19 | 39.6 |
| Provider type |  |  |
| Nurse | 12 | 25 |
| Physician | 12 | 25 |
| Community actor | 24 | 50 |
| Facility type |  |  |
| Public | 38 | 79.2 |
| Specialized | 10 | 20.8 |

**Table 2.**
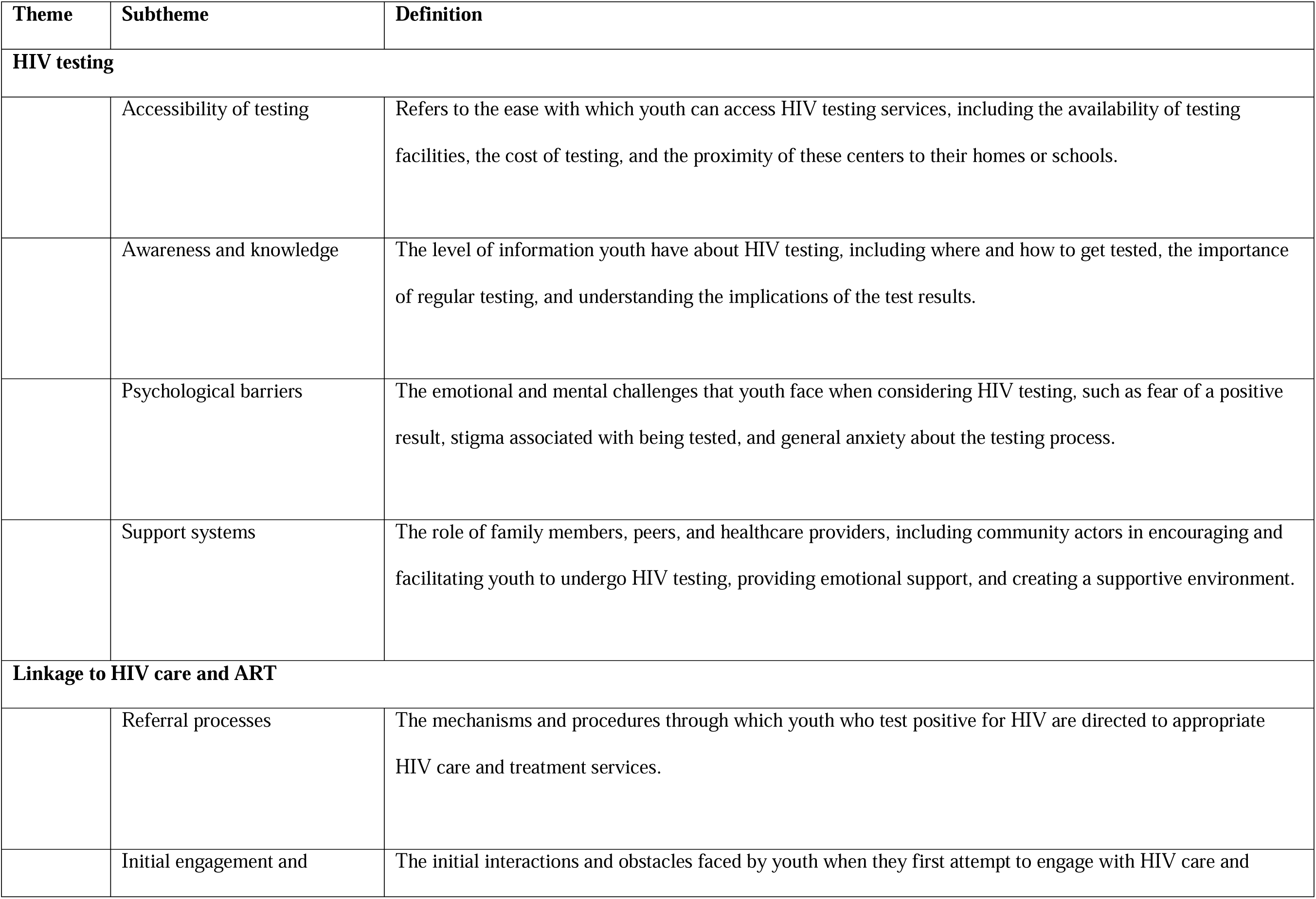
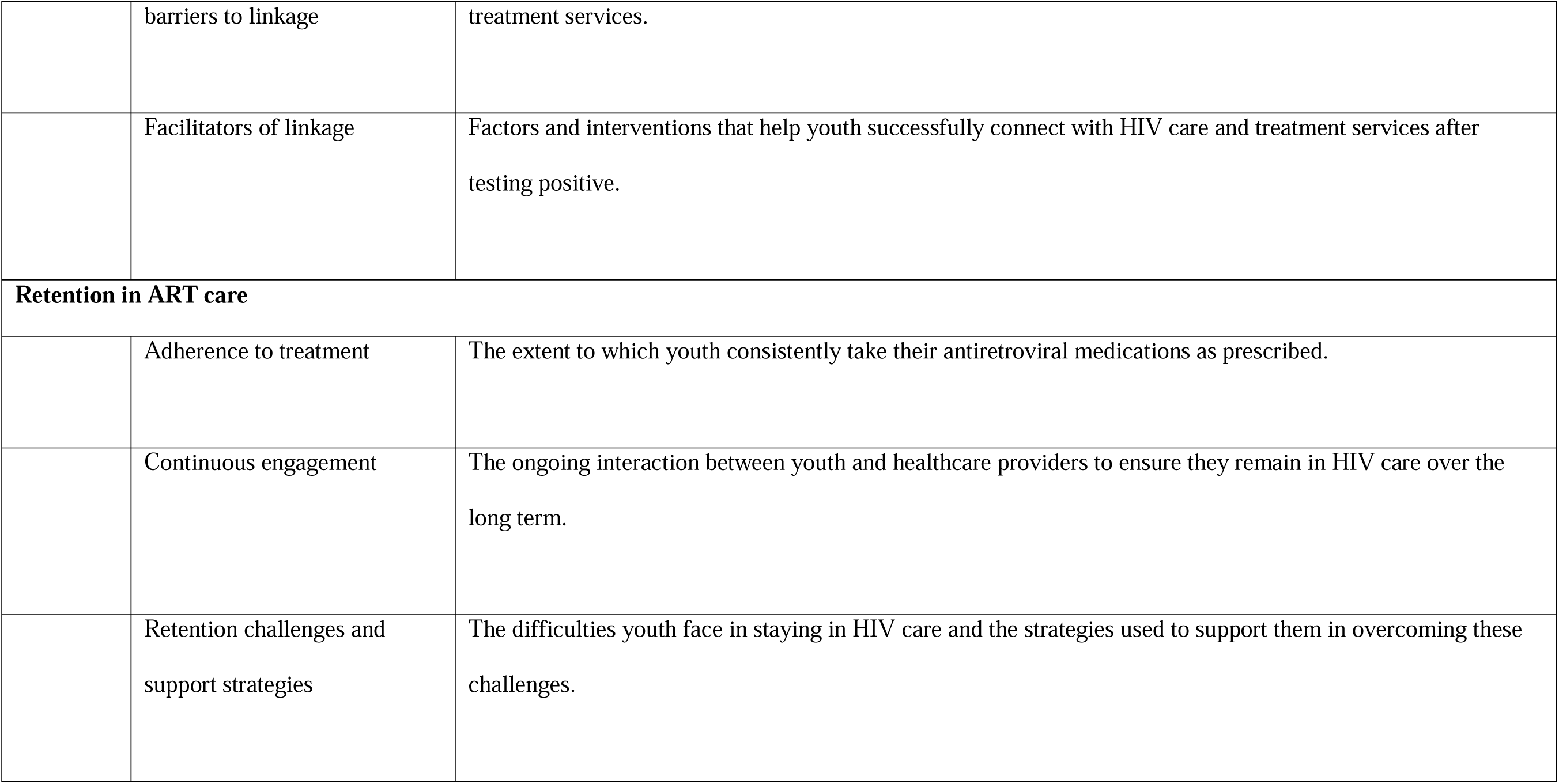
Themes identified through focus group discussions with participants.

Youth, whether HIV-negative individuals or those infected with HIV, face particular challenges accessing effective care and achieving successful treatment outcomes. Fig 1 shows HIV care pathways. This was used to guide the organization and discussion of the generation of themes. Fig 1 has 3 different components, including the (1) UNAIDS HIV/AIDS targets for 2030, (2) system successes, and (3) system failures. The targets for 2030 can be read as follows: 95% of people living with HIV know their HIV status, of which (blue arrow), 95% receive treatment, of which (pink arrow), 95% are virally suppressed. To achieve the global 95-95-95 goals, adopted by United Nations Member States in June 2021, it would require system successes [31–33]. When an adolescent is tested for HIV, two cases can happen: if negative, then this adolescent is linked to counseling and HIV prevention services, including behavioral and/or biomedical strategies. However, if positive, the adolescent follows the HIV care continuum. Finally, the third component showcases the system failures due to challenges at different levels, extensively discussed throughout the manuscript.

**Figure 1.**
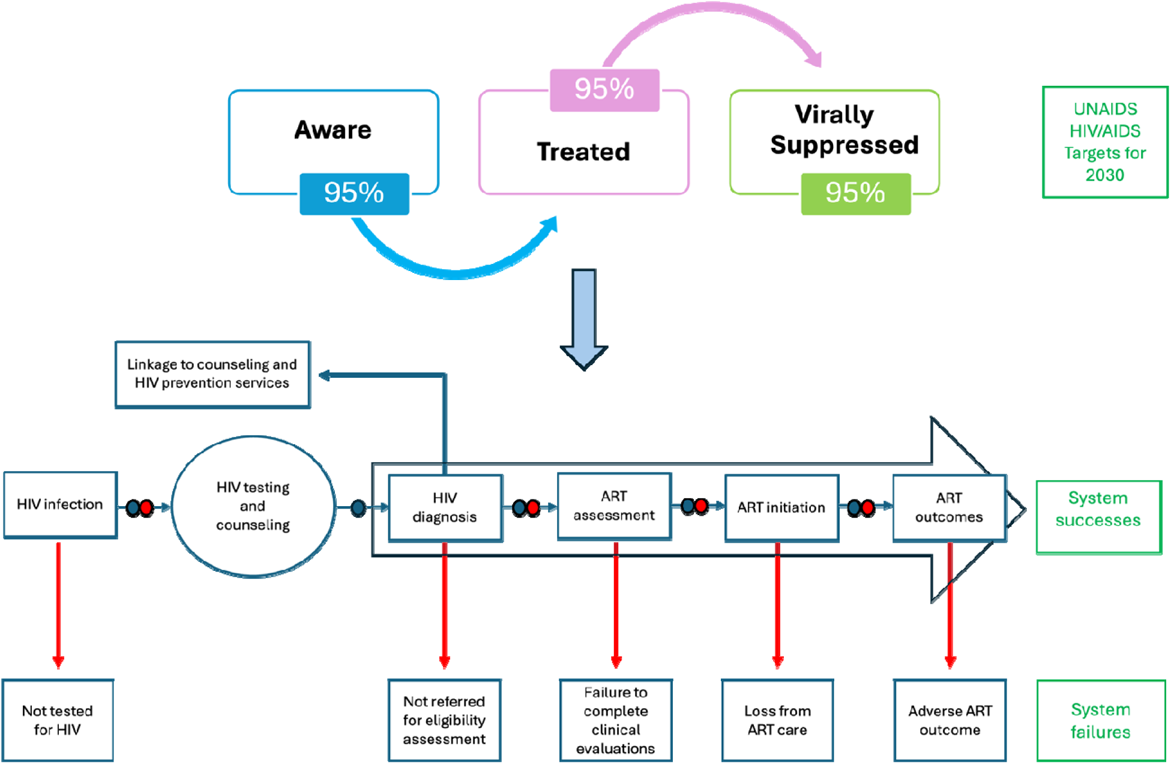
Overall adolescent-focused HIV pathway to care in Chad. **Notes:** In this process, blue and red circles on the arrows indicate the presence of healthcare workers and communit actors, respectively. Figure 3 was informed by the grounded theory data collection, which included in-dept interviews and focus group discussions through a participatory action research. It was further inspired b MacPherson et al. (2012), MacPherson et al. (2015), Herce et al. (2019), UNAIDS (2020); and adapted from a figure by Kranzer et al. (2012).

**Figure 2.**
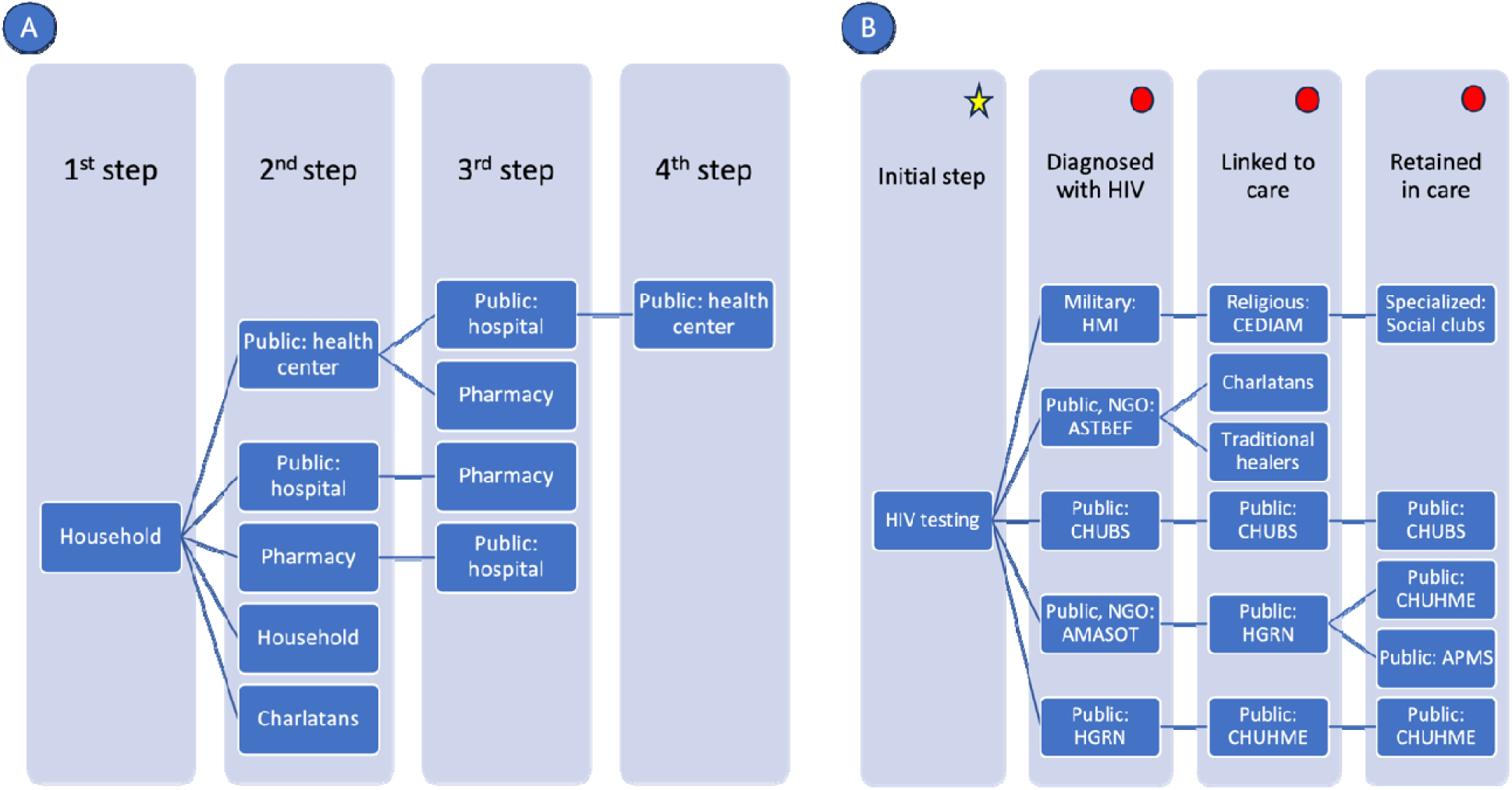
The current pathways to care experienced by A) HIV-negative and B) HIV-positive youth. **Notes:** Public sectors are managed and operated by the Ministry of Health, and include health posts, health centers, and hospitals; Private sectors are privately-run, non-governmental healthcare site; Pharmacy sector is a business selling medications and/or offering consultations for medical complaints, usually for-profit; Traditional healers or those using rituals, herbal remedies, spiritual healing and other methods based on the cultural and spiritual beliefs of their community, usually for-profit. Charlatans or fake doctors are individuals who falsely claim to be qualifie healthcare workers. For figure 4B, yellow star represents HIV-positive youth whose diagnoses often occurred durin routine medical visits or emergencies prompted by recurrent illnesses or family-initiated tests, and red circles represent community actors, including psychosocial counselors, expert patients, mentor moms, and peer educators operating throughout the HIV continuum of care. Additional information on the description of healthcare sectors is in supplementary exhibit 6.

Figures 2A and 2B depict the care pathways for HIV-negative and HIV-positive youth in Chad, respectively. For HIV-negative youth, the journey begins at the household level and progresses through public health centers, hospitals, and pharmacies, with occasional visits to traditional healers and charlatans. This reflects the blend of traditional and modern healthcare practices in Chad. As youth move through these steps, they may repeatedly visit public health centers and hospitals, highlighting the iterative nature of healthcare-seeking and the need for specialized care at higher-level facilities. This pathway illustrates the complex navigation required within the healthcare system for HIV-negative youth. For HIV-positive youth, the process starts with HIV testing at various institutions, followed by diagnosis and linkage to care through military, public, NGO, and religious facilities. The retention in care phase involves specialized social clubs and multiple public health institutions.

#### Theme 1: HIV testing services

##### Subtheme 1: Accessibility of testing

Youth’s access to HIV testing was a critical factor in their engagement with SRH services. Accessibility encompassed the availability of testing facilities, the cost of tests, and the proximity of these centers to their homes or schools. One adolescent described their experience, saying *“I was tested for HIV five months ago at the BABA Moustapha cultural center. It was part of a workshop. Accessing the service was very simple”* (Participant #11, age range 22-24 years). Despite these examples of easy access, the financial aspect remained a barrier for many, as highlighted by another adolescent who noted the high costs associated with hospital visits and the need to seek more affordable options like pharmacies. Furthermore, the lack of free additional testing as healthcare often required multiple screenings were not always covered, making it financially burdensome for youth. For instance, an adolescent shared, *“Going to the hospital for tests costs a lot of money. We young people are looking for shortcuts like going to the pharmacy, which is less expensive”* (Participant 4, age range 20-22 years). Another mentioned, *“Hospitals are expensive for examinations and due to the lack of resources, we can’t do them. If rates can be reduced for us young people, it would be good because we don’t have the financial means”* (Participant #1, age range 20-22 years). These challenges were compounded by issues like reagent shortages, which limited the availability of necessary tests, as noted by a community actor: *“On the financial side: access is voluntary; however, we are dealing with reagent shortages, and this has been the case since August 2023 except for pregnant women (prioritized)“* (Community actor, FGD #1).

Youth approached HIV testing influenced by various factors, including family, social environment, and personal health experiences, navigating fragmented care pathways across multiple healthcare sectors (Fig 2). Regular testing for HIV was mentioned as a common practice among HIV-negative youth, often driven by personal or parental initiative, which highlighted the importance of early detection and periodic testing after risky behaviors such as *“unprotected sex”* (Participant #34, age range 18-19 years). Many youths relied on health centers, hospitals, school programs, family referrals, and health campaigns for testing. For instance, one adolescent shared, *“I am taken by my [parent] for testing. After testing, we went home, and he came back to get the results”* (Participant #26, age range 18-19 years). Similarly, others described straightforward access to testing through school campaigns and cultural center workshops. Healthcare workers and community actors have played pivotal roles in facilitating access to these services. For example, community actors mediated between young people and healthcare facilities, providing information and support to navigate the testing process: *“My role is to help, accompany, and guide them. In my opinion, it is important to raise awareness in churches, schools, and in the neighborhood”* (Community actor, FGD #2). Besides biomedical strategies, behavioral HIV preventive interventions were also employed since service providers offered advice and counseling to HIV-negative youth. Youth reported receiving advice from *“psycho-social counselors, attending physicians”* (Participant #27, age range 18-19 years), and noted that these providers are key sources of information and support.

##### Subtheme 2: Awareness and knowledge

The level of awareness and knowledge about HIV testing among youth significantly influenced their likelihood of getting tested. Youth gathered information from various sources, including *“associations working to combat sexually transmitted diseases”* (Participant #9, age range 15-17 years), *“social networks and educational institutions”* (Participant #23, age range 20-22 years). These sources provided crucial information about where and how to get tested, the importance of regular testing, and understanding the implications of the test results. Some youth received referrals from family members, while others took the initiative to seek testing due to health concerns: *“I did not feel well and went straight to the hospital for testing because I did not want to suffer. The fear of dying was great. I took the test and when I found out it was negative, it prompted me to take preventive measures in the future”* (Participant #4, age range 20-22 years). However, misconceptions and a lack of comprehensive knowledge deterred youth from seeking testing. For instance, one adolescent mentioned, *“Some people think that once you take the test, you will get infected”* (Participant #2, age range 18-19 years). Another added, *“I thought you could only get tested if you were already sick”* (Participant #1, age range 20-22 years). Healthcare workers were instrumental in educating and advising youth about the importance of regular testing and HIV prevention.

##### Subtheme 3: Psychological barriers

Psychological barriers played a significant role in youth’s decision-making regarding HIV testing. Adolescent females often faced higher barriers to accessing HIV testing due to societal norms and stigma associated with sexual health. Females were more likely to encounter negative reactions from healthcare providers and fear of judgement, which discouraged them from seeking testing. As one community actor noted, *“The obstacle is the lack of confidentiality”* and *“family presence is required”* (Community actor, FGD #2). Additionally, some adolescent girls reported being coerced into testing by their parents, sometimes without fully understanding the process or its implications. A healthcare worker mentioned, *“When a teenager presents (always accompanied), if the doctor refers them to us for screening, the child is isolated from the parents to find out if sexual intercourse was protected or not”* (Healthcare worker, FGD #1). In contrast, adolescent males experienced less societal pressure but still faced significant obstacles. Many young men reported challenges related to the convenience of testing facilities and the fear of receiving their status, which deterred them from seeking testing. An adolescent shared, *“It’s not easy. For us students, we have to be on vacation and every time we take the test, we are afraid of being positive”* (Participant #3, age range 22-24 years). This fear could be overwhelming and prevent youth from getting tested, even when they recognized the importance of knowing their status. Despite the fear and stigma associated with HIV testing, some youth still found the courage to get tested, driven by risky behaviors or symptoms, as expressed by: *“The fear is there, but you have to have the strength to go and take the test”* (Participant #3, age range 22-24 years) or *“I do not think it’s easy. You have to be brave*.

*The first day I refused, but once I made love with someone I did not know without protection, I had to find the courage to take the test”* (Participant #2, age range 18-19 years). Addressing these psychological barriers requires creating a supportive and non-judgmental testing environment, providing counseling services to help youth cope with their fears, and normalizing HIV testing as a routine part of healthcare. Healthcare workers and community actors contributed significantly by providing reassurance where youth felt safe and supported during the testing process, as expressed by: *We the psychosocial counselors mediate between young people and the hospital. There are young people who lack information, and some are trying to access these services. So, we are the hyphen, we give them the information”* (Community actor, FGD #1).

##### Subtheme 4: Support systems

Support systems, including family members, peers, and healthcare providers, were crucial in encouraging youth to undergo HIV testing. The presence of a supportive environment can significantly reduce the fear and stigma associated with testing. For example, one adolescent mentioned, *“My [relative] is a nurse so I ask her to take me to the hospital”* (Participant #1, age range 20-22 years), illustrating the importance of having trusted individuals who can facilitate access to testing (Fig 2). Additionally, schools and community organizations played a vital role in providing information and support, helping to create a culture where HIV testing was seen as a responsible and routine health behavior. For HIV-positive youth, diagnoses often occurred during routine medical visits or emergencies prompted by recurrent illnesses or family-initiated tests (Fig 2). An adolescent recounted, *“I had pimples on my body and my [parent] took me to the hospital and after the test, I was diagnosed with HIV”* (Participant #50, age range 15-17 years). Many discovered their status through parental intervention, such as Participant #33 who was screened by their parent, or Participant #32, who was screened due to frequent illnesses. Youth also encountered diagnoses in unexpected medical contexts, like Participant #30, who tested positive during additional tests following surgery. Table 3 shows selected quotes from HIV-positive youth.

**Table 3.**
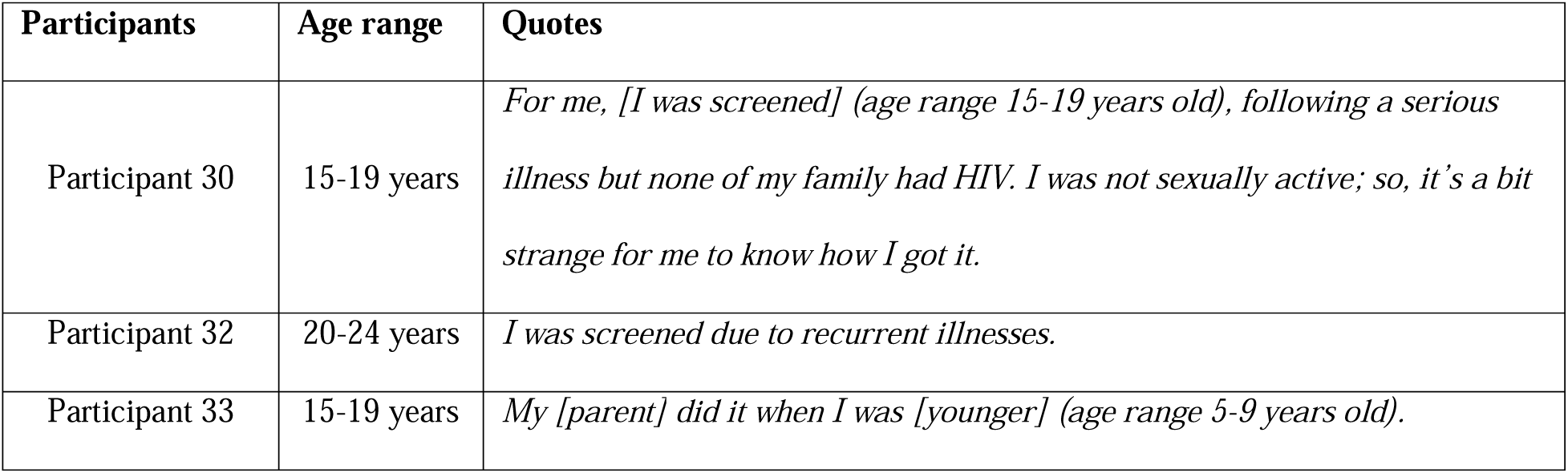
Quotes from HIV-positive youth on diagnoses following HIV testing with family support.

Service providers have been crucial in offering continuous support and follow-up, ensuring that youth remained engaged with the healthcare system. Strengthening these support systems by involving families, peers, and community leaders can enhance youth’s willingness to get tested and ensure they receive the necessary support throughout the process. Thus, while these barriers were significant, the support systems and personal determination of youth played crucial roles in their decision to undergo HIV testing.

#### Theme 2: Linkage to HIV care and ART

##### Subtheme 1: Referral processes

This process was shaped by various factors, from initial referral processes to the barriers and facilitators youth encountered. Effective referrals and supportive healthcare professionals were crucial in helping youth transition from testing to treatment. For instance, one adolescent recounted, *“The psychosocial counselor gave me advice, which helped me to accept treatment”* (Participant #44, age range 20-22 years). Healthcare workers played a vital role in this stage by providing clear guidance and support, ensuring that youth understood the importance of immediate engagement with care, as expressed by, *“It’s not the end of the world. You are going to take ARTs and be like other kids. You should come in for the viral load tests in 3-4 months, so if it’s undetectable, you can’t transmit the virus to another person anymore, and you are going to do whatever other people do too”* (Healthcare worker, FGD #2).

##### Subtheme 2: Initial engagement

Youth’s first experiences with HIV care were important in determining their long-term engagement with treatment. Positive initial interactions with healthcare providers could significantly influence their decisions to seek care post-diagnosis. One adolescent highlighted this importance, saying, *“It’s difficult at first, but once trust is established between the patient and doctor, everything becomes easy”* (Participant #37, age range 22-24 years). Healthcare workers often provided emotional support and reassurance during this critical period. For example, one healthcare worker mentioned, *“My role is to see the adolescent living with HIV with a smile. It’s very important for me to listen, advise, guide, and lead the person to access his diagnosis”* (Healthcare worker, FGD #3). Additionally, a community health worker said, *“When they learn that they are infected they believe that life is over. But when we tell them that with treatment, they will have HIV-negative children and then, they find hope again”* (Community actor, FGD #1). This supportive approach helped youth overcome initial fears and uncertainties about treatment.

However, despite these efforts, youth faced significant barriers to linkage to HIV care and ART. Financial constraints were a major obstacle, as noted by an adolescent who said, *“I have no money to pay for medication“* (Participant #44, age range 20-22 years). Due to this situation, some youths were forced to seek non-traditional care options such as charlatans and traditional healers, further endangering their health conditions (Fig 2). Logistical challenges, such as transportation issues and the fear of stigma, also hindered effective linkage. Community actors highlighted the importance of confidentiality and trust, with one noting, *“Dealing with teenagers is difficult and there needs to be an emphasis on confidentiality”* (Community actor, FGD #1). Some youth found it easier to discuss their health with community actors such as peers rather than healthcare workers due to the perceived lack of confidentiality. This was further discussed by a healthcare worker who noted, *“Youth often fear the judgment and lack of confidentiality from healthcare workers”* (Healthcare worker, FDG #1). A community actor explained, *“These youth are not wrong because some youths are not kind and expose children”* (Community actor, FGD #1), which emphasized the need to create a supportive and stigma-free healthcare environment.

##### Subtheme 3: Facilitators of linkage

Youth’s preferences for certain facilities over others, despite geographic proximity, were influenced by several factors. They often favored specialized HIV health facilities and social clubs over public hospitals due to the quality of care, the reception, and the supportive environment provided. As one adolescent noted, in specialized establishments, *“care is assured and their products are effective because they are subsidized“* whereas in public hospitals, *“care is not total and the welcome is not as good“* (Participant #26, age range 18-19 years). Specialized facilities and social clubs were perceived to offer more comprehensive and empathetic care, free from stigma and discrimination, which was crucial for youth managing sensitive health issues like HIV. This sentiment was echoed by another adolescent who mentioned that in clubs, *“we feel good”* (Participant #45, age range 20-22 years) whereas in public hospitals, the staff sometimes *“neglect and stigmatize us”* (Participant #44, age range 20-22 years).

Additionally, the availability of trained staff, confidentiality, and effective communication in specialized centers contributed to youth’s preferences. Social clubs, in particular, offer a space for open dialogue and guidance, making them attractive to young people seeking both medical and psychosocial support. As one adolescent described, *“we share everything in words; they encourage us to go to the hospital to get tested and to use the services“* (Participant #4, age range 20-22 years). These facilities also provided a sense of community and emotional support, which public hospitals often lack. Although healthcare workers emphasized the need for continuous education and support, with one stating, *“We must advise them and support them as much as possible”* (Healthcare worker, FGD #2), the specialized facilities and social clubs stood out because they offered tailored environments that prioritized confidentiality, empathy, and comprehensive care. This combination of factors made them particularly appealing to youth who required both medical and psychosocial support in managing their health.

#### Theme 3: Retention in ART care

##### Subtheme 1: Adherence to treatment

Youth’s commitment to staying on ART was often influenced by their understanding on the importance of consistent medication intake and managing side effects. One adolescent expressed their determination, stating, *“I agreed to continue ART to avoid transmitting HIV to others and to stay healthy”* (Participant #50, age range 15-17 years). Healthcare workers played a crucial role in this process by providing clear and consistent information about the benefits of adherence and helping manage any side effects that may arise. For example, a healthcare worker noted the importance of therapeutic education, stating, *“It is important to remain clear about the examples of therapeutic education”* (Healthcare worker, FGD #3). This clarity helped youth understand the critical nature of their treatment regimen and reinforced their commitment to maintaining it.

##### Subtheme 2: Continuous engagement

Retention in ART care also involved regular follow-ups with healthcare providers and continuous engagement with healthcare services. Frequent interactions with healthcare providers helped reinforce the importance of adherence and provide ongoing support. One adolescent reflected on their routine, mentioning, *“When you feel like you’re taking a risk during sexual intercourse, you must be aware and take precautions, including regular follow-ups and adherence to ART”* (Participant #34, age range 18-19 years), illustrating the continuous engagement needed. Healthcare workers emphasized the need to spend adequate time with patients to build trust and ensure they understand their treatment. One healthcare worker stated, *“The patient must always have a little bit of time with the doctor after the first consultation because he is the one who accompanies him through the process”* (Healthcare worker, FGD #4). This ongoing relationship between healthcare providers and youth is essential for maintaining adherence and addressing any issues that may arise during treatment.

##### Subtheme 3: Retention challenges and support strategies

In previous steps of the HIV care continuum, barriers such as stigma, financial constraints and logistical challenges were identified for HIV testing as well as during the process of linkage to HIV care and ART, respectively. Despite the support from healthcare providers, financial difficulties, transportation issues, and experiences of stigma or discrimination further hindered retention in ART care (Fig 2). For example, logistical issues such as transportation made regular clinic visits challenging. One adolescent shared, *“It’s difficult to access care because of travel”* (Participant #34, age range 18-19 years). Another example involved a teenager who preferred to travel a longer distance to a different health center to avoid stigma, *“There is this teenager who lives in a [city] and prefers to go to a health center in [another city] for the pills despite the referral proposal I made to him and this for the simple reason that his [relative] works at this health center close to him“* (Community actor, FGD #1). Youth have advocated for tailored services to address these challenges, expressing a desire for services that cater specifically to their needs, including tailored hours that accommodate their schedules. For instance, one adolescent suggested, *“We need special centers for teenagers to make them feel comfortable, with hours that fit our school and work schedules“* (Participant #8, age range 15-17 years). This sentiment was echoed by community actors who recognized the need for flexible service hours: *“It would be important to create special structures with everything in it, including tailored hours that allow youth to access care without disrupting their daily routines”* (Community actor, FGD #1).

Youth often stopped treatment when they started feeling better, which disrupted their long-term care. Healthcare workers have responded to these needs by providing longer drug stocks to youth who have earned their trust, which helps to maintain continuity in their treatment. One community actor explained, *“We call them (by phone) if they don’t keep the appointments. We’re going to pick them up (move). At first, they are put on ARVs and when they get better, they leave and only come back when there is a complication”* (Community actor, FGD #1). Another one shared, *“When we gain their trust and they become more attached to us than their own parents, we can trust them with 3 to 6 months of drug stocks“* (Community actor, FGD #1). This approach not only addressed the logistical challenges but also helped ensure that youth remain engaged in their ART regimen.

## Discussion

Findings from this study highlight the significant barriers and facilitators in the HIV care continuum for youth in Chad. The ideal pathway to SRH and HIV care for youth necessitates an adolescent-centered design that prioritizes customized accessibility, responsiveness of care, clinical effectiveness, and tailored service delivery. This approach would significantly improve the HIV care continuum, including HIV testing, linkage to care, and retention in care, by addressing the unique needs and challenges faced by youth.

Accessibility remains a fundamental barrier to HIV testing and care for youth. Our study revealed gender disparities, especially among adolescent females facing higher barriers to accessing HIV testing due to societal norms and stigma. However, adolescent males faced barriers related to the convenience of testing facilities and the fear of receiving a positive status. Like other studies, stigma was also identified as a main barrier to care and treatment adherence among youth living with HIV in many settings in sub-Saharan Africa [4, 34–36]. Youth further emphasized the importance of integrating testing services into familiar and convenient locations, especially due to stigma. Integrating HIV testing into a multi-disease approach, as suggested by youth has been shown to increase efficiency while reducing stigma in SSA, especially among men [37, 38]. Financial and logistical barriers significantly hindered youth’s access to HIV testing and care services. Many youth face challenges such as high transportation costs, long travel distances, and limited availability of specialized HIV care facilities. To address these issues, youth and service providers suggested the establishment of more adolescent-specific centers in close proximity. These centers should be strategically located to minimize travel distances and associated costs, making it easier for youth to access the services they need. By reducing financial and logistical barriers, and providing services within their communities, these centers can play a pivotal role in improving HIV care engagement and outcomes for youth.

The responsiveness of care is crucial for maintaining youth’s engagement in HIV services. Youth reported that negative interactions with healthcare providers, long waiting times, and lack of confidentiality deterred them from seeking and continuing care. Similar challenges have been reported by studies, highlighting these health-system and structural barriers, further complicating youth’s engagement in HIV care. However, solutions exist that can mitigate these challenges. Building on the creation of specific centers for youth, these spaces need to be safe and supportive environments, delivering screening services, psychosocial support, and adhering to adolescent-specific treatment protocols. Additionally, youth highlighted the need for healthcare providers to be more understanding and respectful, which can be facilitated through targeted training programs for healthcare workers on adolescent-friendly practices. Research has shown that engaging youth in the design of HIV care interventions significantly enhances their engagement and acceptance of services. Furthermore, Akama et al. (2023) demonstrated that youth-led initiatives can improve care engagement by making services more responsive to youth’s needs [39].

Ensuring clinical effectiveness involves providing comprehensive and continuous care tailored to the specific needs of youth. Continuous, and specialized training for healthcare workers is crucial, especially in how to deliver adolescent-specific services. This training should focus on developing skills in communication, confidentiality, and the management of adolescent health issues to ensure that healthcare providers can offer empathetic, respectful, and effective care. Recent research has demonstrated the effectiveness of such approaches in improving linkage to and retention in care [40, 41]. Furthermore, in adolescent-centered design, the care provided is based on the best available evidence and is specifically tailored to the unique developmental and psychosocial aspects of adolescent health. This involves using interventions that have been proven effective for this age group and ensuring that healthcare providers are trained in adolescent health dynamics. Clinical effectiveness is enhanced by continuity of care and by integrating feedback from youth to improve and adapt clinical practices and treatment protocols. For instance, studies have shown that youth-friendly service models and human-centered design approaches significantly improve linkage to care and retention rates among youth [42, 43].

Finally, tailored service delivery requires a holistic approach that integrates biomedical, behavioral, psychosocial, and structural perspectives. Youth emphasized the positive impact of having peer educators and psycho-social counselors who provided both information and emotional support, making the healthcare experience more relatable and less intimidating. Recent research supports this integrated approach, showing that peer-led interventions and the inclusion of psycho-social support within youth-friendly spaces significantly improve access and engagement, as well as enhance linkage to care and retention rates among youth [42–46]. Additionally, service providers advocated for the development of community-based programs that involve family and community leaders to reduce stigma and support youth’s adherence to treatment. For example, Willis et al. (2019) demonstrated that interventions involving community adolescent treatment supporters (CATS) significantly improved adherence, retention, and psychosocial well-being among HIV-positive youth in Zimbabwe [47]. Further studies have shown that these community-based and multi-faceted interventions are crucial in addressing the complex needs of youth and improving their overall engagement in HIV care [48]. By incorporating these strategies, healthcare systems can create a more supportive and effective environment that promotes better health outcomes for youth.

Despite the valuable insights gained from this study, several limitations should be acknowledged. Although the study involved 23 FGDs with a diverse sample of youth and service providers, the participants were recruited from specific regions and contexts, which may not fully represent the broader adolescent population in Chad or similar settings. Future research should consider expanding the geographic scope to include rural areas and other provinces to capture a wider range of experiences and perspectives. Second, the reliance on self-reported data presents a potential bias, as participants might have provided socially desirable responses or may not accurately recall their experiences related to HIV care and ART. This could lead to an underestimation or overestimation of certain barriers or facilitators discussed during the FGDs. Triangulating self-reported data with observations could enhance the reliability of the findings. Third, language and translation issues pose another limitation. While the thematic analysis was conducted in the original languages (French and Arabic) to preserve linguistic and cultural nuances, the subsequent translation into English might have resulted in subtle shifts in meaning or loss of context. Lastly, the study’s cross-sectional design captures a snapshot of participants’ experiences at a single point in time, limiting the ability to observe changes over time or assess long-term outcomes related to HIV care and ART adherence. Prospective qualitative studies would be beneficial in understanding how experiences and challenges faced by youth evolve over time.

## Conclusion

Significant progress toward ending the HIV epidemic among youth requires redesigning healthcare services in Chad to be more adolescent-friendly and responsive to their specific needs. This involves integrating biomedical, behavioral, psychosocial, and structural perspectives into a comprehensive approach to HIV care. Tailored interventions focused on reducing stigma, enhancing accessibility, and providing continuous psychosocial support are essential. Continuous research and collaboration with local communities and stakeholders are crucial to developing effective strategies that support youth in navigating the healthcare system and achieving better health outcomes.

## Supporting information

COREQ Checklist

## Acknowledgments

We are grateful for the collaboration and support received from the Blue Cross Chad, Rose Service-Learning Fellowship, Fostering Diversity in HIV Research Program, Tessa Jowell Fellowship for Doctoral Research, Gamble Scholarship, and Chad’s Prime Minister’s Office. We thank the study participations for their willingness to share their experiences and perspectives, which has provided an essential foundation for our research. The study received funding from the Tessa Jowell Fellowship for Doctoral Research and Chad’s Prime Minister’s Office.

## Abbreviations

ART: antiretroviral therapy
AIDS: acquired immunodeficiency syndrome
CDC: Centers for Disease Control and Prevention
CGT: constructivist grounded theory
CNLS: National AIDS Control Council
DHS: demographic and health surveys
FCAS: fragile and conflict-affected settings
FGD: focus group discussion
HIV: human immunodeficiency virus
LGBTQ+: lesbian, gay, bisexual, transgender, and questioning (or queer)
NGO: non-governmental organization
PAR: participatory action research
RNTAP+: National Executive Secretary of the Chadian National Network of Associations of People Living with HIV
SRH: sexual and reproductive health
SSA: Sub-Saharan Africa
UN: United Nations
UNAIDS: Joint United Nations Programme on HIV/AIDS
UNICEF: United Nations International Children’s Emergency Fund
WHO: World Health Organization
YLHIV: youth living with HIV

## Data availability statement

The data underlying this article cannot be shared publicly due to ethical restrictions. When applying for ethical approval, we did not specify that the data would be made publicly available in a repository. As part of the written and verbal consent, we assured participants that all data would be confidential and access to the recordings would be restricted to the research team. We did specify that “some of their words” may be used in reporting the findings of the study (which we have done within the manuscript as non-identifiable quotes), however, to make all raw data publicly available will be a serious breach to the rights of ethical of participants given did not consent to this.

