## Supplementary material for "Rethinking HIV care for youth: Insights from qualitative research with youth in Chad": S1 File

**S1 File: Stages and processes of the constructivist grounded approach employed for parent study to enable a youth sensemaking outcome on youth-centered decision making in HIV care in Chad.**

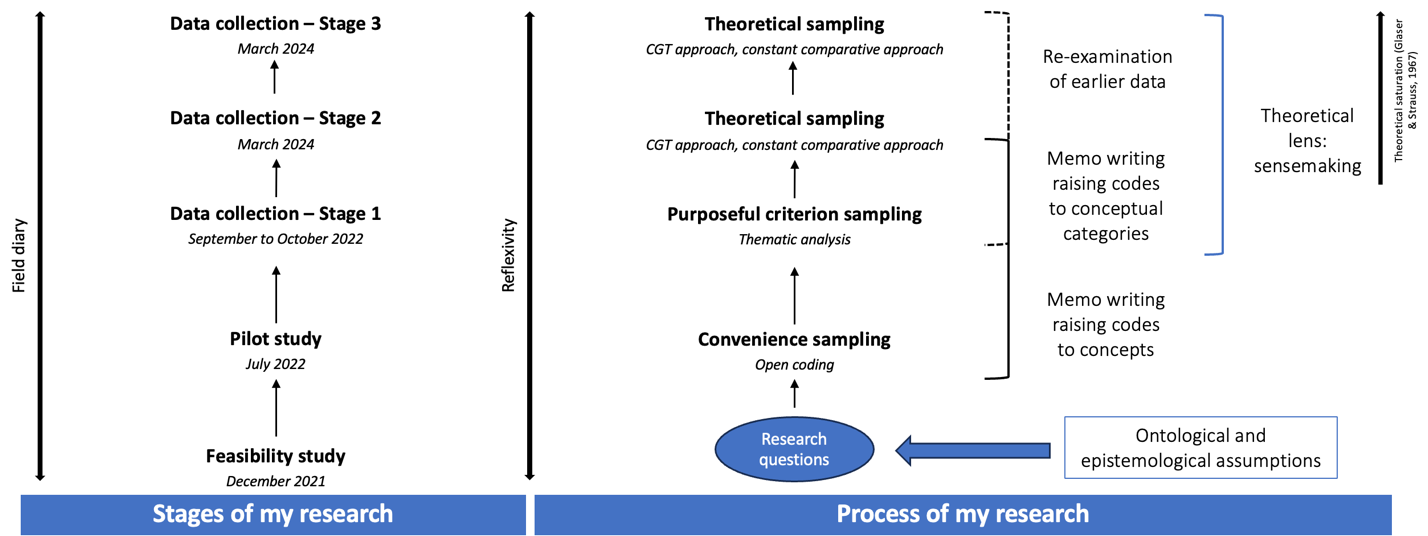

For the study from which the data come from, we followed a constructivist grounded theory (CGT) methodology, with sensemaking as a theoretical lens [21, 22]. The CGT approach was deemed necessary to answer our research questions as we sought to construct a theory explaining the complex interactions influencing how youth access and make sense of the information they have, and how this in turn, shapes how they access, navigate, and use available SRH and HIV services. Prior to stage 1, we conducted feasibility and pilot studies in December 2021 and July 2022 respectively. These were done to obtain insights into the study setting and type of participants needed for data collection stage 1. For stage 1, which took place between September and October 2022, we used a purposeful sample criterion sampling method. In stage 1, we sought to assess the contextual factors and mechanisms that influence access to information and health services, as well as understand the youth’s experience of HIV supportive norms in Chad [49]. For stages 2 and 3, we focused on the youth as well as service providers (i.e., healthcare workers and community actors), respectively. Due to the limited evidence on youth’s access to SRH and HIV care in Chad, this study was done to address this gap by offering insights into the unique challenges and challenges faced by Chadian youth in accessing these services. For all stages, theoretical sampling was used. This resulted in the final model: Theoretical model of empowerment of the youth in health decision-making through adaptive sensemaking.

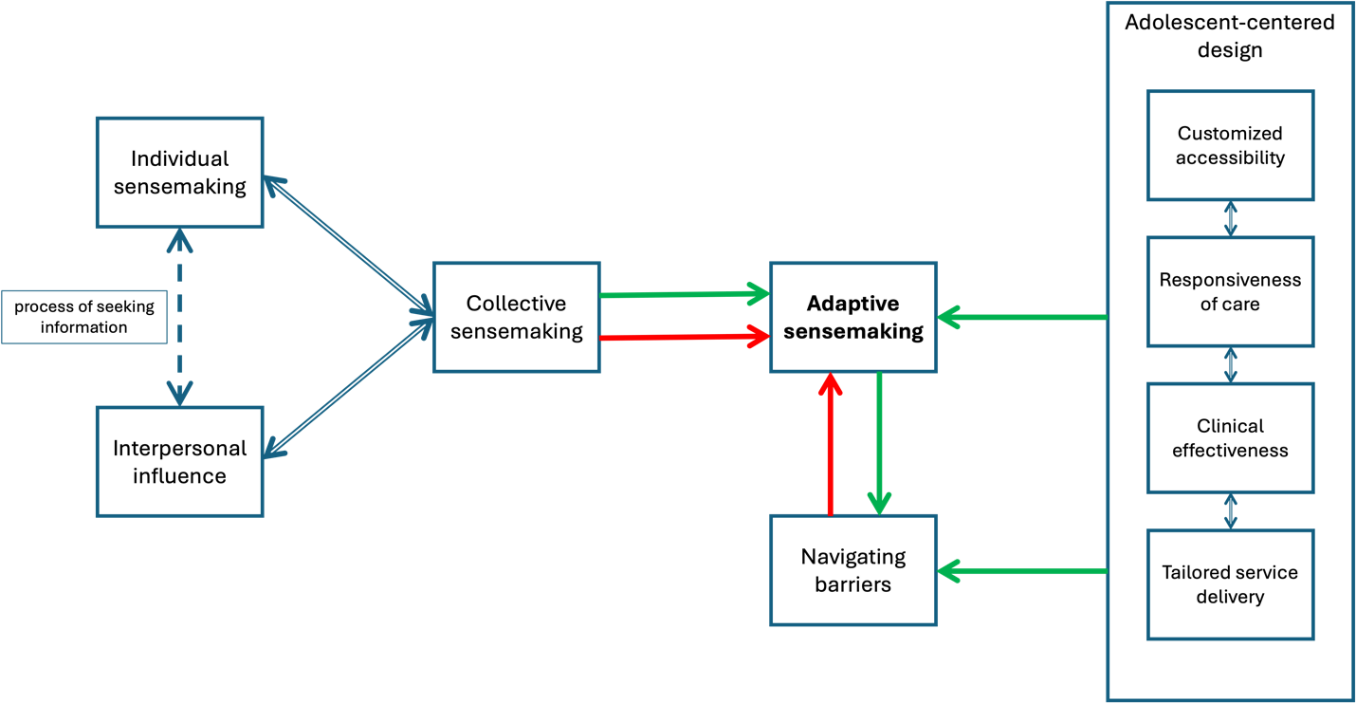

Notes: Taken from Bedingar et al., 2024a^[1]^.

This model rests on three key components, including collective sensemaking, navigating obstacles, and enhancing accessibility and quality through designs centered around adolescents. This theory proposes that to enable young people to effectively make decisions about their health, especially in areas such as SRH and HIV care, it is crucial to adopt an adaptive sensemaking approach. This approach should recognize the interconnectedness of personal experiences, social influences, and broader systemic factors. By prioritizing these elements, our aim is to help youth better navigate their healthcare journeys. Ultimately, this approach is expected to bolster their interaction with healthcare services, promote their utilization, improve treatment adherence, and offer insights for refining healthcare policies and services (Bedingar et al., 2024a).

1. Unpublished work.
