## Supplementary material for "Rethinking HIV care for youth: Insights from qualitative research with youth in Chad": S2 File

**S2 File: Flowchart of sub-divided participants in each group.**

***HIV negative group: male (N = 16)***

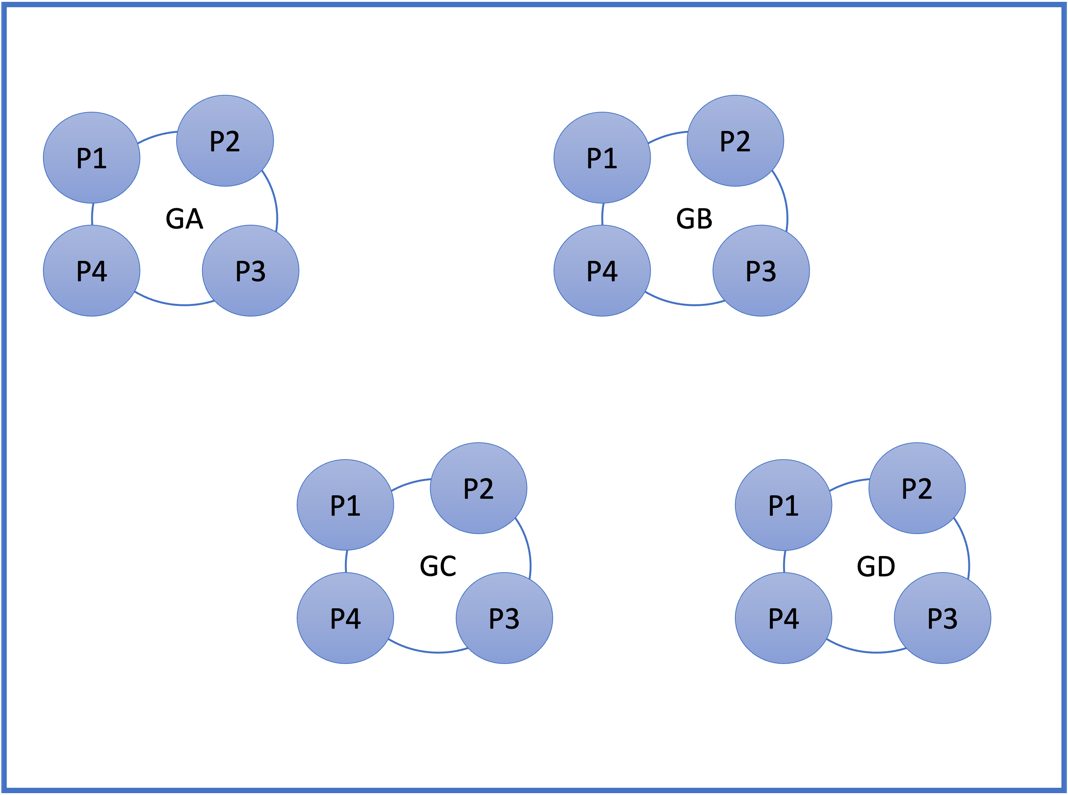

***HIV negative group: female (N = 13)***

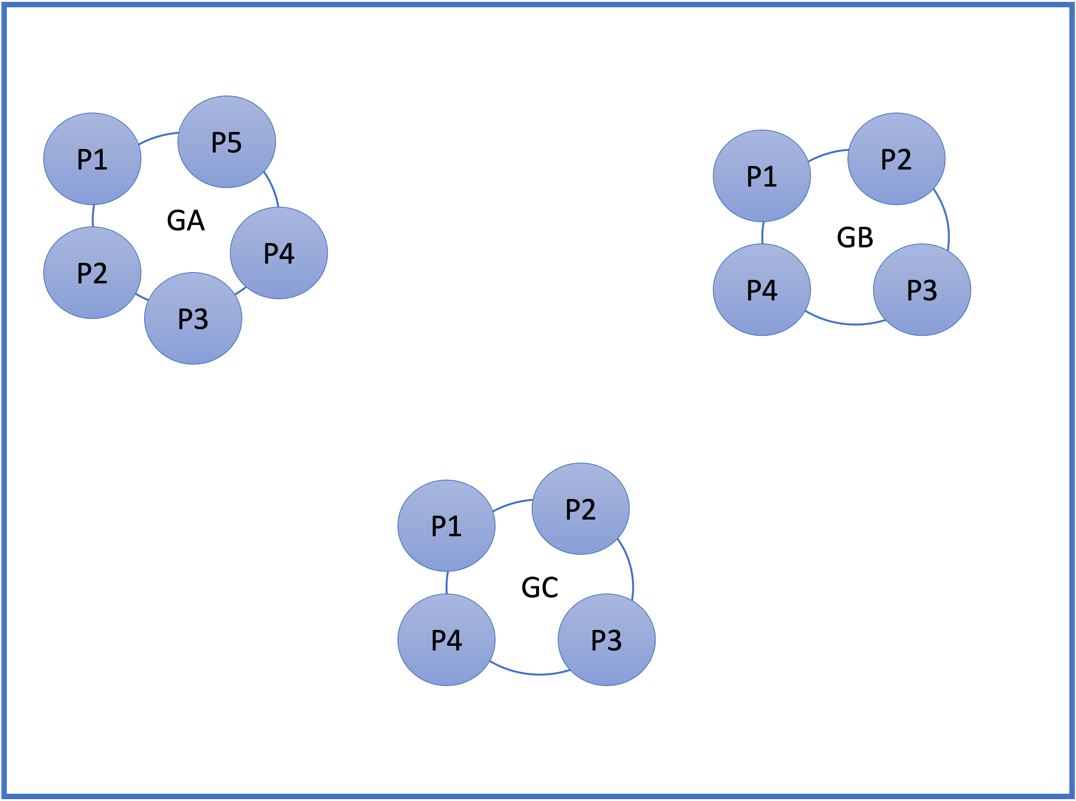

***HIV positive group: male (N = 12)***

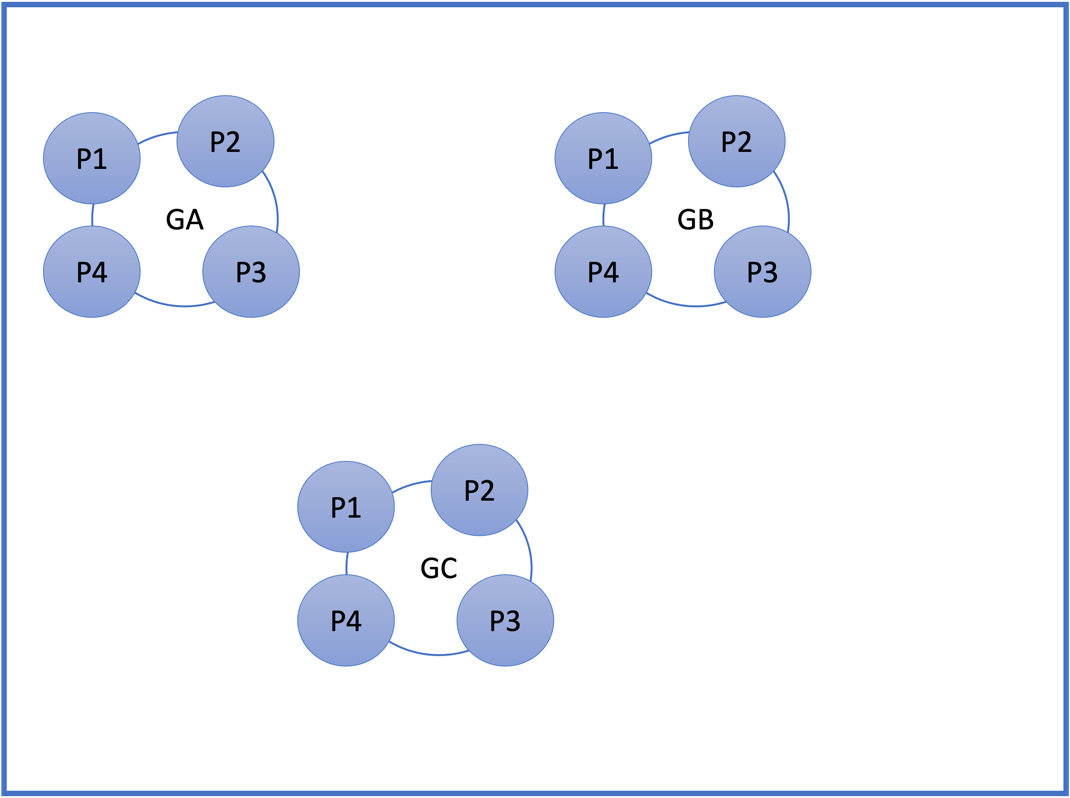

***HIV positive group: female (N = 11)***

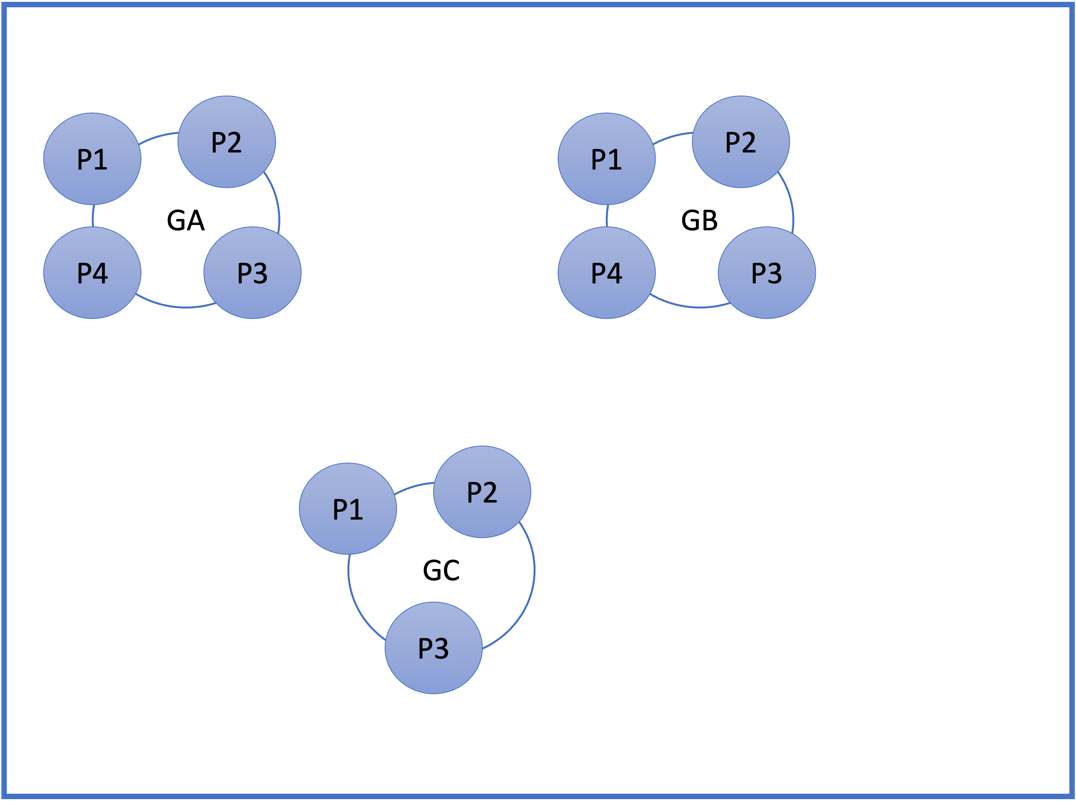
