## Supplementary material for "Rethinking HIV care for youth: Insights from qualitative research with youth in Chad": S3 File

**S3 File: Focus group discussion topic guide for youth.**

BRIEF BACKGROUND: Since 2010, there is a 48% decrease in new HIV infections. Despite this decrease, youth are disproportionately affected by the epidemic. According to UNAIDS, 26.3% of new HIV infections were in youth and young people (aged 15-24 years) in 2022. Despite all the progress made, young women continue to suffer disproportionately from the HIV epidemic as they have HIV infection rates four times (0.42%) as high as young men (0.1%), accounting for 19.5% of all new HIV infections. This presents an opportunity to understand how youth access and ‘make sense’ of the information they have, how they make decisions to seek for help, and how this in turn, shapes how they navigate and use available services. Insights from this study will allow the design, implementation and scale-up of more effective and youth-centric and friendly interventions in Chad.

**Topic Guide**

Hello [name]!

We are conducting a study to learn about sexual and reproductive health and HIV access to information and services among the youth in Chad. You are being asked to participate in this focus group discussion/in-depth interview because your insights and experiences are relevant to this study. If you take part in this study, you will be asked to participate in a workshop where you will be part of in-group activities from 7.00am to 5.00pm on a given day. The day will be divided into sessions for each theme, which will take 90-120 minutes per theme. Please refer to the consent form for more details.

Consent Process

1. **Please introduce yourselves within your own group**
2. **Theme 1: Making sense of sexual and reproductive health (SRH) and HIV care**
   a. If I mention SRH and/or HIV care, what would you think of? What does SRH and/or HIV care mean to you?
   b. What services do you see as being included under SRH and HIV care? Where do you get most of your information?
   c. Why would you use SRH and HIV care? In what circumstances?
3. ***Theme 1 Group activity:***
   a. If you have ever used SRH and/or HIV care, please can you walk me through the process of making choices or if you have not, imagine…
   b. How would you decide what to do? How do you choose what service to use?
      i. [for each decision], probe for what were the influencing factors? Was this factor an enabler or a barrier to receiving the service, in what way?
4. **Theme 2: Expectations, beliefs, and knowledge about SRH and HIV?**
   a. What services are available to you? Perceptions of alternative avenues of SRH and HIV care?
   b. What do you think the differences are between public vs. specialized HIV health facilities vs. social clubs?
   c. What are the sources of advice or information about SRH and HIV care services?
5. ***Theme 2 Group activity:***
   a. Do you think SRH and/or HIV care services could be improved for young people? If so, how?
   b. Please draw your care paths:
      i. Your current care path, taking into consideration [reason to go, choice of facility, ability to reach facility, did you seek the service alone/with someone, waiting for the health worker, interactions with health workers, service received, follow up]
      ii. [Review it] and look at how you would have changed it for it to be ideal.
6. **Theme 3: Previous experiences of using SRH and/or HIV care**
   a. Have you used SRH and/or HIV care services before? How easy or difficult do you think it was to access care?
      i. Probe for: How convenient are different sorts of facilities [public vs. private vs. specialized vs. confessional/religious]?
   b. Your experiences of using SRH and/or HIV care in recent years – timeline of symptoms, decisions and satisfaction with care received?
      i. Probe: What are the alternatives to this care?
      ii. Probe: How has this experience shaped your thoughts about future health seeking?
7. **Theme 4: Prioritizing influencing choices**
   a. Reflect on specific instances of using SRH and/or HIV care, the circumstances surrounding these and the factors which influenced these decisions.
   b. Reflect on times when you did not use SRH and/or HIV care, and on what influenced decisions not use these services.
8. ***Theme 4 Group activity:***
   a. Reflect on a time you needed either SRH or HIV services and you went to get it or if you have not, then imagine…on each [index] card write down one reason for going to get the services. After writing on as desired, prioritize factors/reasons from the most to the least influential. Then, write down the meaning of your card piles. Finally, discuss piles with your group.
   b. Reflect on time you needed either SRH or HIV services and you decided not to go…on each [index] card write down one reason for going to get the services. After writing on as desired, prioritize factors/reasons from the most to the least influential. Then, write down the meaning of your card piles. Finally, discuss piles with your group.
9. **Theme 5: The burden of access to healthcare – navigating and accessing care**
   a. What is your experience with the availability of services at the local level?
   b. For some people, the personal work of managing their health can be…
      i. Emotionally difficult?
      ii. Financially difficult?

Is there anything else you would like to share with us about the issues we have explored together today?

Thank you for your time.
