## Supplementary material for "Rethinking HIV care for youth: Insights from qualitative research with youth in Chad": S4 File

**S4 File: Focus group discussion topic guide for healthcare workers.**

BRIEF BACKGROUND: Since 2010, there is a 48% decrease in new HIV infections. Despite this decrease, youth are disproportionately affected by the epidemic. According to UNAIDS, 26.3% of new HIV infections were in youth and young people (aged 15-24 years) in 2022. Despite all the progress made, young women continue to suffer disproportionately from the HIV epidemic as they have HIV infection rates four times (0.42%) as high as young men (0.1%), accounting for 19.5% of all new HIV infections. Using previously collected data, the youth mentioned that they refused to seek care due to the quality of care received and power dynamics exerted by healthcare workers when asked about their experience. This presents an opportunity to focus on healthcare workers and understand how they support and provide an enabling environment for the youth to seek care. Insights from this study will allow the design, implementation and scale-up of more effective and youth-centric and friendly interventions in Chad.

**Topic Guide**

Hello [name]!

We are conducting a study to learn about sexual and reproductive health and HIV access to information and services among the youth in Chad, especially how healthcare workers provide an enabling environment for the youth to seek care. You are being asked to participate in this focus group discussion/in-depth interview because your insights and experiences are relevant to this study. If you take part in this study, you will be asked to participate in a workshop where you will be part of in-group activities from 7.00am to 1.00pm on a given day. As the day will be divided into sessions for each theme, which will take 90-120 minutes per theme. Please refer to the consent form for more details.

Consent Process

1. **Setting the scene**
   a. Please introduce yourselves within your own group
2. **Theme 1: Making sense of their role as a healthcare worker/service provider**
   a. How do you perceive your role as a healthcare worker?
   b. What impact (health outcomes) do you want to have in your patients’ lives?
      i. Probe: for youth, specifically.
3. **Theme 2: Defining quality and adolescent-friendly healthcare**
   a. As it relates to youth and young people, if you hear me say “quality and adolescent-friendly healthcare services”, what would you think of?
   b. If you were to define quality, what would you say? If you were to define adolescent-friendly services, what would you say?
      i. Probe: Are they different? Similar? Or what would you say?
4. **Theme 3: Prioritizing influencing factors**
   a. Reflect on a recent time you had to provide SRH or HIV care [e.g., testing, contraception, pregnancy, symptoms, etc.] to an adolescent or young person, what did you do for each of these instances below. For each instance, please explain the influencing factors (why did you act in such way). Was this factor an enabler or a barrier to providing the service described, in what way?
      i. Your knowledge and skills as a service provider
      ii. The level of respect you showed to your patient
      iii. The clarity of your explanations
      iv. The involvement of your patient as much as they wanted to be in decisions about their care
      v. The amount of time spent with your patient
      vi. The courtesy and helpfulness you showed to your patient
   b. Go back to the examples you mentioned you gave, and reflect on whether you could change it; if yes, then tell us, what you would have done.
5. ***Theme 3 Group activity:***
   a. As a group, based on your understanding of quality and adolescent-friendly healthcare, reflect on what you consider as *“creating a conducive environment for the youth to seek care”*, then write on each [index] card factors or elements that you think will enable it. After writing on as desired, prioritize these cards from the most to the least important factor. Then, write down the meaning of your card piles. Finally, discuss piles with your group.
   b. [Read the definition of quality of care], then ask them to re-sort the card piles
6. **Theme 4: Making sense of quality and adolescent-friendly healthcare**

**System competence**

- Safety
- Prevention and detection
- Continuity and integrity
- Timely action
- Population health management

**Care competence**

- Provider competence
- Timely action
- Systematic assessment
- Correct diagnosis
- Appropriate treatment
- Counseling and education
- Referral

**Foundations**

- Equipment
- Medicines
- Data

**User experience**

- **Customer service**
  - Short wait time
  - Ease of use
  - Choice of provider
  - Patient voice and values
  - Affordability
- **Respect**
  - Dignity
  - Privacy
  - Clear communication
  - Non-discrimination
  - Autonomy
  - Confidentiality

7. ***Theme 4 Group activity:***
   a. In groups, please put these quality-of-care domains in either of these buckets. After putting them in buckets [refer to the processes of care: care competence, system competence, user experience, foundations], please pick them up one by one and explain why this specific bucket, and what does it [quality-of-care domain] mean to you as it relates to SRH and HIV care for youth. You are also welcome to leave any of the QoC domain you deem not important.
