## Supplementary material for "Rethinking HIV care for youth: Insights from qualitative research with youth in Chad": S5 File

**S5 File: Focus group discussion topic guide for community actors.**

BRIEF BACKGROUND: Since 2010, there is a 48% decrease in new HIV infections. Despite this decrease, youth are disproportionately affected by the epidemic. According to UNAIDS, 26.3% of new HIV infections were in youth and young people (aged 15-24 years) in 2022. Despite all the progress made, young women continue to suffer disproportionately from the HIV epidemic as they have HIV infection rates four times (0.42%) as high as young men (0.1%), accounting for 19.5% of all new HIV infections. Using previously collected data, the youth mentioned that they refused to seek care due to the quality of care received and power dynamics exerted by healthcare workers when asked about their experience. This presents an opportunity to focus on healthcare workers and understand how they support and provide an enabling environment for the youth to seek care. Insights from this study will allow the design, implementation and scale-up of more effective and youth-centric and friendly interventions in Chad.

**Topic Guide**

Hello [name]!

We are conducting a study to learn about sexual and reproductive health and HIV access to information and services among the youth in Chad, especially how healthcare workers provide an enabling environment for the youth to seek care. You are being asked to participate in this focus group discussion because your insights and experiences are relevant to this study. If you take part in this study, you will be asked to participate in a workshop where you will be part of in-group activities from 7.00am to 1.00pm on a given day. As the day will be divided into sessions for each theme, which will take 90-120 minutes per theme. Please refer to the consent form for more details.

Consent Process

1. **Setting the scene**
   a. Please introduce yourselves within your own group
2. **Theme 1: Making sense of their role as a community actor [according to the type, including peer educators, mentor moms, expert patients, and psycho-social counselors]**
   a. How do you perceive your role as a [type of community actor]?
      i. Probe: What impact do you want to have in the youth’s lives?
   b. Explain how do you interact with youth?
3. **Theme 2: Previous experience with youth**
   a. Based on your experience, describe the challenges and barriers that youth face when it comes to seeking HIV care.
   b. For peer educators (including for sex workers and MSM):
      i. Due to their status as key populations, how do you influence their sensemaking processes for the following actions:
         1. Getting tested
         2. If positive, taking ARVs
         3. Staying on ARVs (follow-up)
   c. For mentor moms:
      i. For adolescent mothers (ages 15-24), how do you influence their sensemaking processes for the following actions:
         1. Getting tested
         2. If positive, taking ARVs
         3. Staying on ARVs (follow-up)
   d. For expert patients:
      i. For adolescent (ages 15-24), how do you influence their sensemaking processes for the following actions:
         1. Getting tested
         2. If positive, taking ARVs
         3. Staying on ARVs (follow-up)
   e. For psycho-social counselors:
      i. For adolescent (ages 15-24), how do you influence their sensemaking processes for the following actions:
         1. Getting tested
         2. If positive, taking ARVs
         3. Staying on ARVs (follow-up)
4. ***Theme 3: Examining the pathways of care***
   a. Reflect on a recent time you had to support an adolescent or young person, then please draw the care path of the adolescent, including your role at each point and discuss.
      i. Probe: What did you do?
      ii. Probe: When faced with a challenge, what factors did you use to influence their decisions?
   b. In your own opinion, was this care path drawn ideal? If not, what would you add to this care path to make it ideal?
5. **Theme 4: Creating a conducive environment for youth**
   a. Discuss specific strategies employed to create a safe and supportive environment for youth. Include what’s missing now to make it a reality.
   b. Provide examples of working interventions that you have used to reduce stigma and improve access to care.
   c. How do you adapt your communication support strategies to resonate with young individuals?
