## Supplementary material for "Rethinking HIV care for youth: Insights from qualitative research with youth in Chad": S1 Table

**S1 Table: Themes identified through focus group discussions with participants.**

| **Theme** | **Subtheme** | **Definition** | **Sample quotes** |
| --- | --- | --- | --- |
| **HIV testing** | | | |
|  | Accessibility of testing | Refers to the ease with which youth can access HIV testing services, including the availability of testing facilities, the cost of testing, and the proximity of these centers to their homes or schools. | *I was tested for HIV five months ago at the BABA Moustapha cultural center. It was as part of a workshop. Accessing the service was very simple.* |
|  | Awareness and knowledge | The level of information youth have about HIV testing, including where and how to get tested, the importance of regular testing, and understanding the implications of the test results. | *I didn't know much about HIV testing until my school organized a health awareness day. Now, I understand why it's important to get tested regularly.* |
|  | Psychological barriers | The emotional and mental challenges that youth face when considering HIV testing, such as fear of a positive result, stigma associated with being tested, and general anxiety about the testing process. | *I was really scared to get tested because I was afraid of what the result might be. What if it was positive? I didn't want people to look at me differently.* |
|  | Support systems | The role of family members, peers, and healthcare providers, including community actors in encouraging and facilitating youth to undergo HIV testing, providing emotional support, and creating a supportive environment. | *My older sister went with me to get tested. She talked to me about it and made me feel less scared. The nurse was also very kind and supportive.* |
| **Linkage to HIV care and ART** | | | |
|  | Referral processes | The mechanisms and procedures through which youth who test positive for HIV are directed to appropriate HIV care and treatment services. | *After my test came back positive, the clinic immediately referred me to a specialist who could start my treatment right away.* |
|  | Initial engagement and barriers to linkage | The initial interactions and obstacles faced by youth when they first attempt to engage with HIV care and treatment services. | *I had a hard time finding the clinic they referred me to. It was far from my home, and I didn't have the money for transportation.* |
|  | Facilitators of linkage | Factors and interventions that help youth successfully connect with HIV care and treatment services after testing positive. | *The community health worker came to my house and helped me understand what I needed to do next. They even arranged transport for my first visit.* |
| **Retention in ART care** | | | |
|  | Adherence to treatment | The extent to which youth consistently take their antiretroviral medications as prescribed. | *Taking my medicine every day was tough at first, but the support group I joined really helped me stay on track.* |
|  | Continuous engagement | The ongoing interaction between youth and healthcare providers to ensure they remain in HIV care over the long term. | *The clinic checks in with me regularly to see how I'm doing and if I need any help with my treatment.* |
|  | Retention challenges and support strategies | The difficulties youth face in staying in HIV care and the strategies used to support them in overcoming these challenges. | *Sometimes I want to give up because it's hard to stay motivated, but my counselor always finds a way to encourage me and keep me going.* |
