## Supplementary material for "Rethinking HIV care for youth: Insights from qualitative research with youth in Chad": S2 Table

**S2 Table: Healthcare sectors identified for the current pathways to care for HIV-negative youth.**

| **Sector** | **Description** |
| --- | --- |
| Public: Ministry of Health | Health posts, health centers and hospitals open to general public, managed and operated by the Ministry of Health. |
| Specialized for HIV | Public, but specialized in HIV care. |
| Private | Privately run, non-government healthcare site. |
| Pharmacy | Business selling medications and/or offering consultations for medical complaints, usually for-profit. |
| Witch doctors or traditional healers | Rituals, herbal remedies, spiritual healing, and other methods based on the cultural and spiritual beliefs of their community, usually for-profit. |
| Charlatans or marabout | Individuals who falsely claim to be qualified healthcare professionals. Locally called “Dr Choukou”, they offer medical advice, treatments, and procedures without having the necessary qualifications, training, or licenses. |
| Household | Treatment or care from a member of participants’ social network (family member, friend, neighbor, etc.). |
