## Supplementary material for "Rethinking HIV care for youth: Insights from qualitative research with youth in Chad": S3 Table

**S3 Table: Healthcare sectors identified for the current pathways to care for HIV-positive youth.**

| **Sector** | **Type** | **Abbreviation** | **Name** | **Activities** |
| --- | --- | --- | --- | --- |
| Public | University Hospital Center | CHUBS  CHUHME | University Hospital Le Bon Samaritain  University Hospital for Mothers and Children | Involved in medical management, from HIV testing to medical examinations, and ART. |
| Public | National Reference Hospital | HGRN | National Reference General Hospital | Involved in medical management, from HIV testing to medical examinations, and ART. |
| Public | NGO | ASTBEF | Chadian Association for Family Well-Being | Runs static clinics and mobile operations to offer voluntary counselling and testing for HIV. |
| Public | NGO | AMASOT | Social Marketing Association in Chad | Works in AIDS prevention activities, through social marketing programs for behavior change. |
| Public | Public | APMS | Psycho-medico social support center | Complements information, education, and communication activities aimed at behavioral change, the medical management of opportunistic infections, access to ART and biological monitoring, right through the care of survivors. |
| Religious | NGO | CEDIAM | Center for Education, Information and Patient Support | Bases its activities on awareness raising on HIV/AIDS, HIV voluntary screening, medical and psychological support, and school tutoring for HIV orphans. |
| Military | Military Hospital | HMI | Military Training Hospital | The hospital’s anti-HIV/AIDS unit offers free, comprehensive treatment of the infection, from screening to ART, including viral load. |
| Charlatans or marabout |  |  |  | Individuals who falsely claim to be qualified healthcare professionals. |
